# Relevance of prediction scores derived from the SARS-CoV-2 first wave, in the UK COVID-19 second wave, for early discharge, severity and mortality: a PREDICT COVID UK prospective observational cohort study

**DOI:** 10.1101/2021.06.09.21258602

**Authors:** Hakim Ghani, Alessio Navarra, Phyoe K Pyae, Harry Mitchell, William Evans, Rigers Cama, Michael Shaw, Ben Critchlow, Tejal Vaghela, Miriam Schechter, Nazril Nordin, Andrew Barlow, Rama Vancheeswaran

**Author notes:** **Corresponding author:** Hakim Ghani, Respiratory Department, West Hertfordshire Hospitals NHS Trust, Vicarage Road, Hertfordshire WD18 0HB, United Kingdom. Joint first authors (Hakim Ghani, Alessio Navarra).

## Abstract

**Objective:** Prospectively validate two prognostic scores, pre-hospitalisation (SOARS) and hospitalised mortality prediction (4C Mortality Score), derived from the coronavirus disease 2019 (COVID-19) first wave, in the evolving second wave with prevalent B.1.1.7 and parent D614 severe acute respiratory syndrome coronavirus 2 (SARS-CoV-2) variants, in two large United Kingdom (UK) cohorts.

**Design:** Prospective observational cohort study of SOARS and 4C Mortality Score in PREDICT (single site) and multi-site ISARIC (International Severe Acute Respiratory and Emerging Infections Consortium) cohorts.

**Setting:** Protocol-based data collection in UK COVID-19 second wave, between October 2020 and January 2021, from PREDICT and ISARIC cohorts.

**Participants:** 1383 from single site PREDICT cohort and 20,595 from multi-site ISARIC cohort.

**Main outcome measures:** Relevance of SOARS and 4C Mortality Score derived from the COVID-19 first wave, determining in-hospital mortality and safe discharge in the UK COVID-19 second wave.

**Results:** Data from 1383 patients (median age 67y, IQR 52-82; mortality 24.7%) in the PREDICT and 20,595 patients from the ISARIC (mortality 19.4%) cohorts showed both SOARS and 4C Mortality Score remained relevant despite the B.1.1.7 variant and treatment advances. SOARS had AUC of 0.8 and 0.74, while 4C Mortality Score had an AUC of 0.83 and 0.91 for hospital mortality, in the PREDICT and ISARIC cohorts respectively, therefore effective in evaluating both safe discharge and in-hospital mortality. 19.3% (231/1195, PREDICT cohort) and 16.7% (2550/14992, ISARIC cohort) with a SOARS of 0-1 were potential candidates for home discharge to a virtual hospital (VH) model. SOARS score implementation resulted in low re-admission rates, 11.8% (27/229), and low mortality, 0.9% (2/229), in the VH pathway. Use is still suboptimal to prevent admission, as 8.1% in the PREDICT cohort and 9.5% in the ISARIC cohort were admitted despite SOARS score of 0-1.

**Conclusion:** SOARS and 4C Mortality Score remains valid, providing accurate prognostication despite evolving viral subtype and treatment advances, which have altered mortality. Both scores are easily implemented within urgent care pathways with a scope for admission avoidance. They remain safe and relevant to their purpose, transforming complex clinical presentations into tangible numbers, aiding objective decision making.

**Trial registration:** NHS HRA registration and REC approval (20/HRA/2344, IRAS ID 283888).

## INTRODUCTION

The overwhelming burden of the severe acute respiratory syndrome coronavirus 2 (SARS-CoV-2) pandemic on global healthcare is well reported with severe ongoing impact in lower resource healthcare systems^1-4^. Over 160 million people have been infected with global mortality exceeding 3.5 million^5^. Infections continue to escalate when restrictions are lifted, with a mutating virus adapting to ensure infectivity, evading both vaccination and host adaptive immunity^6,7^. The pandemic is evolving, documented by heterogeneity in outcomes between the first and second waves. Differing unadjusted mortality and case fatality rate are recorded, higher in South Africa, Belarus, and Russia but contrastingly lower in Japan, Italy, United Kingdom (UK), United States of America (USA), Spain and Sweden in the second wave^8-14^. India and Nepal, affected severely in the lethal second wave exemplifies this variation where the first wave was more sanguine^15^.

The UK variant, B.1.1.7, accounted for 58% to 83% of all second wave UK hospitalised cases with increased infectivity but reduced mortality compared to the parent D614G^16,17^. This started in early September 2020, peaking on the 1st of January 2021 and prior to any significant vaccination effort^18^. A younger (60y vs 62y), less frail (12.8% vs 22.8%), more obese (29.1% vs 24.6%), and more female (47.3% vs 41.8%) case population in the second wave was noted in a small London study^17^. Spain and Japan noted similar demographic changes with reduced mortality but without documented SARS-CoV-2 variants^11,13^. Age, the most significant predictor of mortality in early reports, was less strongly predictive of case fatality with substantial reduction in nursing home mortality in many first world countries, although it remained the same in Denmark and Norway but increased in Australia^12^. South Africa noted a higher mortality in the second wave thought to be due to a combination of an overloaded healthcare system, less restrictive public health measures, under-reported mortality in the first wave and the B.1.351 subtype^9,19^.

The changing behaviour noted in coronavirus disease 2019 (COVID-19) considered due to evolving SARS-CoV-2 subtypes, socio-economic healthcare responses and/or varying host-viral interactions, suggests difficulties in prognostication^4,6,7,20-22^. An early scramble to produce COVID-19 severity scores to help transform complex clinical pictures into objective decision aids resulted in a plethora with varying efficacy^23^. Of these, the 4C Mortality Score validated in a large multi-site UK cohort using International Severe Acute Respiratory and Emerging Infections Consortium World Health Organization Clinical Characterisation Protocol UK (ISARIC) data from first wave hospitalised SARS-CoV-2 patients, is widely used^24^. SOARS, a rapid clinical score with multi-site validation including the ISARIC cohort, is a peer reviewed model that enables safe, reliable, and expedient discharge on presentation to any urgent care area, making it invaluable during peak pressures^25^. Scores stratifying mortality and deterioration are all based on the first wave of the pandemic without updates advocated by the evolving pandemic.

This study updates the performance of both the 4C Mortality Score and SOARS scores in the UK COVID-19 second wave, characterised by the predominant B.1.1.7 variant, in both the derivation and multi-site cohorts: PREDICT and ISARIC. Prospective validation particularly of the early discharge score (SOARS) remains vital for reliable healthcare resource planning, enabling resumption of usual services in countries with lower infection rates^1,2^. We predict that subsequent waves will infect lower-risk patients, who will benefit most from safe triage for home discharge or to the supportive virtual hospital (VH)^26,27^. Safe and rapid triage to home care cannot be over-emphasised as the pandemic threatens the developing world with a huge scarcity of hospital beds^4^.

## METHODS

### Study design and characteristics of cohorts

Adults 18 years and older who tested positive for SARS-CoV-2 nucleic acid by real-time reverse transcriptase PCR (rRT-PCR) between 1^st^ October 2020 to 25^th^ January 2021 (defined as the UK COVID-19 second wave) after presenting to the Emergency Department (ED) at West Hertfordshire Hospitals NHS Trust, were prospectively recruited (PREDICT second wave cohort). Comparison data representing the first wave (March to May 2020) with similar inclusion criteria was previously collected^25^. Baseline clinical characteristics and investigations were collected according to a pre-specified protocol in a National Health Service Health Research Authority (NHS HRA) and Research Ethics Committee (REC) approved study (20/HRA/2344, IRAS ID 283888)^25^. Patients were either discharged, referred to the Virtual Hospital (VH) for outpatient monitoring or admitted to the hospital^27^. We also received an additional 20,595 UK COVID-19 second wave data from ISARIC (ISARIC second wave cohort) to determine the performance of SOARS and 4C Mortality Score in the UK COVID-19 second wave.

### Laboratory, physiologic and radiographic data

All laboratory tests were performed as part of routine clinical care. Nasopharyngeal mucosal swabs for rRT-PCR were tested in a recognised UK Public Health England laboratory. Recorded baseline vital observations included all the parameters recommended by the previously described scores: SOARS and 4C Mortality Score^24,25^. Chest radiographs (CXR) acquired in ED were collated and scored at the end of the recruitment period by two independent respiratory physicians and verified if discordant by a respiratory radiologist. Each lung field was divided into upper, middle, and lower zones, and one point was scored for each zone affected.

### Location and level of care

After presentation to the ED at West Hertfordshire Hospitals NHS Trust, the clinical course was protocol and pathway driven as per NHS HRA and REC 20/HRA/2344, IRAS ID 283888 (Supplement 1)^25^. Patients who were clinically judged to have mild infection were referred to the VH for subsequent monitoring. Patients who were admitted but did not require additional respiratory support beyond supplemental oxygen were managed on designated medical wards. Where clinically indicated, high-flow nasal oxygen (HFNO) or continuous positive airway pressure (CPAP) were provided on respiratory level-2 wards supported by respiratory physicians or in the intensive care unit (ICU). Intubation and mechanical ventilation were undertaken in the ICU. All hospital admitted patients were treated with Dexamethasone as per NICE guidance. Remdesivir and Tocilizumab were offered where clinically indicated, according to current available evidence^28^.

### Patient and public involvement

As this study was initiated early in the pandemic (March 2020), it was neither feasible nor appropriate to involve patient or public participation in the design, recruitment, conduct, writing or dissemination of this study. This contrasted with the virtual hospital care design in the previous pre-specified protocol used for this study with patient engagement^25,27^. However, for admitted patients, as per the local hospital protocol for escalation of care management plans, a religious advisor and a non-executive non-clinical director were consulted during multidisciplinary meetings.

### Validation of scores derived from the first wave in the second wave cohorts

The primary outcome of the study was in-hospital mortality, a measure used to prospectively evaluate published COVID-19 scores^24,25^. Variables considered relevant in the SOARS and 4C Mortality Score studies with numerically relevant odds ratio (OR) or a p-value of <0.05 were included in the final analysis. All variables required for the 4C Mortality Score (8 variables: sex, age, number of comorbidities, Glasgow coma scale, respiratory rate (RR), oxygen saturation (SpO2), urea and C-reactive protein (CRP)) and SOARS (5 variables: SpO2, obesity, age, RR and history of stroke) were collected and prospectively evaluated in the time period defined previously^24,25^. The ability to discriminate for in-hospital mortality was assessed by the area under the receiver operating characteristic (AUC). We then externally validated scores with the additional 20,595 UK COVID-19 second wave data received from ISARIC.

### Statistical analysis

Categorical variables were expressed as frequency (%), with significance determined by the Pearson’s χ2 test. Continuous variables were expressed as median (IQR) or mean (SD) and analysed by the t-test, Kruskal-Wallis or Mann-Whitney U test, as appropriate. P values were adjusted by Bonferroni correction. A p value of <0.05 was considered statistically significant. Receiving operating characteristics (ROC) curves for SOARS and 4C Mortality Score were constructed by multiple linear regression of the variables included in the respective score. This was done both in our population (PREDICT) and in the ISARIC dataset comprising of 20,595 patients. Patients with missing values in one or more of the variables included in the regression model have been excluded from the calculation. All statistical analyses including risk modelling calculations were performed using GraphPad PRISM statistics software (GraphPad, San Diego, USA) and R statistical language.

## RESULTS

### Comparison of characteristics between waves

1383 patients (53.4% male) confirmed as SARS-CoV-2 rRT-PCR-positive were prospectively recruited over the 14-week study period (from 1^st^ October 2020 to 25^th^ January 2021) from West Hertfordshire Hospitals NHS Trust, UK (PREDICT second wave cohort). The baseline characteristics of the patients in both waves are compared in Table 1.

**Table 1.**
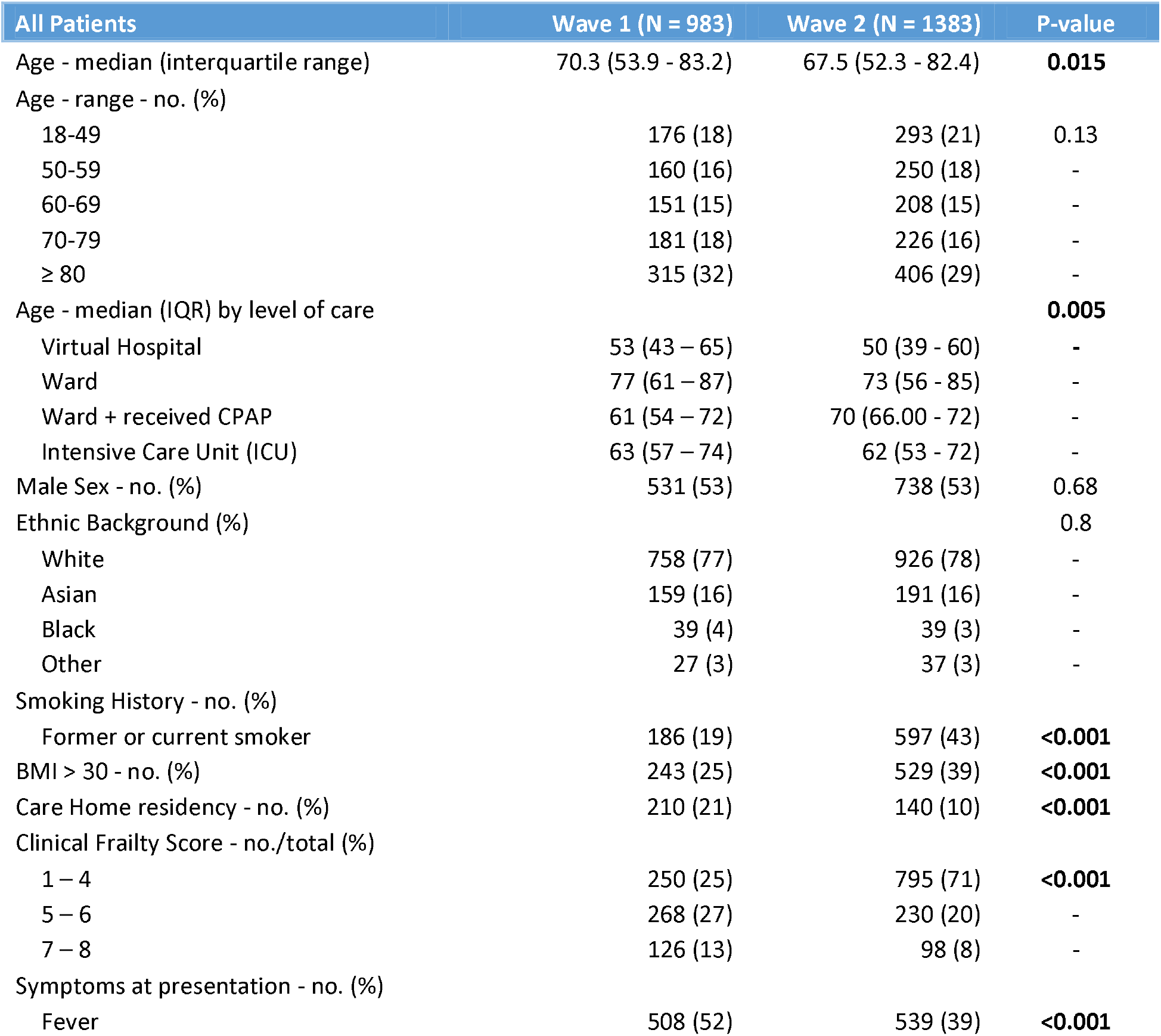
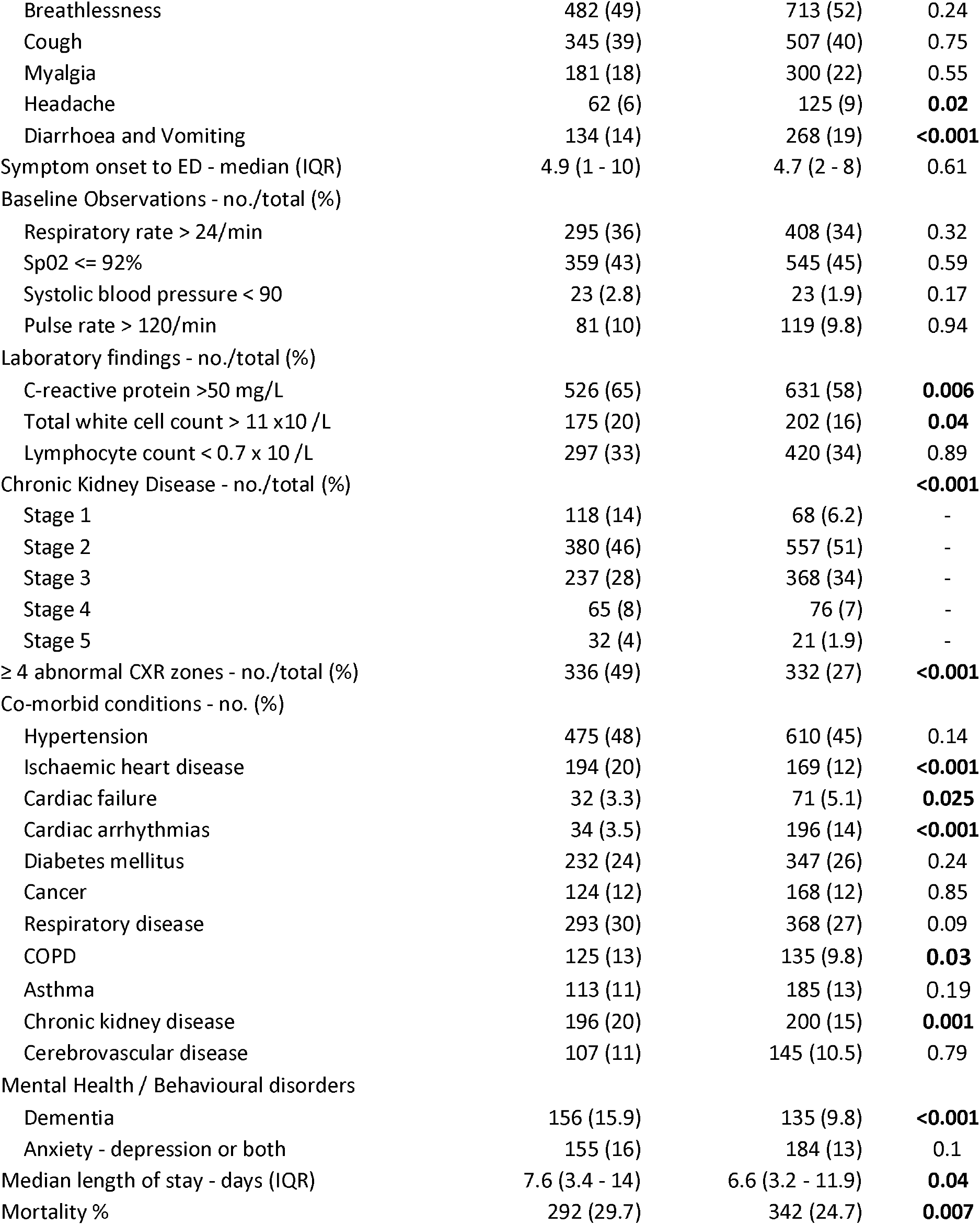
Baseline characteristic of first and second wave (PREDICT cohort)

When comparing the first wave (March to May 2020) and second wave (October 2021 to Jan 2021), there was a significant decrease in median age, 70.3y (IQR: 53.9 – 83.2) vs 67.5y (IQR: 52.3 – 82.4), with no difference in male sex (53% vs 53%) and ethnicity, (77% White, 16% Asian, 4% Black vs 78% White, 16% Asian, 3% Black). The second wave noted increased patients with smoking history (19% vs 43%) and BMI >30 (25% vs 39%). Patients were significantly less frail, with reduced number of Clinical Frailty Score (CFS) >5 (40% vs 28%), less likely to have dementia (15.9% vs 9.8%) and care home residence patients halved (21% vs 10%) in the second wave. A lower prevalence of cardiovascular, respiratory, and renal diseases was noted in the second wave, with significantly lower number of patients with ischaemic heart disease (20% vs 12%, P < 0.001) and chronic obstructive pulmonary disease (13% vs 9.8%, P = 0.03). There was similar prevalence of diabetes (24% vs 26%), cancer (12% vs 12%) and asthma (11% vs 13%)

Presenting symptoms of fever was less prevalent but gastrointestinal symptoms (diarrhea and vomiting) and headache were more prevalent in the second wave. No differences were noted in respiratory or hemodynamic observations on presentation: respiratory rate, oxygen saturation, SF ratio, blood pressure and heart rate. Blood investigations in ED noted reduced patients with CRP >50 mg/L (65% vs 58%) and WCC > 11 × 10^9/L (20% vs 16%), but no difference in lymphopenia in the second wave. Chest radiology (CXR) was more severe in the first wave with significantly higher number of patients with severe CXR scores of >4 zones (49% vs 27%).

Dexamethasone was only given to all admissions in the second wave as per updated NICE guideline in comparison to the first wave where dexamethasone was not prescribed. Remdesivir and Tocilizumab, 10.7% and 2% respectively, were given only in the second wave according to current available recommendation^28^. Mortality and intubation in patients on Tocilizumab and/or Remdesivir compared with age matched second wave patients did not show any change in outcomes. The two waves noted similar time from symptom onset to presentation, but reduced length of stay in hospital and mortality from 29.7% to 24.7%. Characteristics that predicted mortality in the PREDICT second wave cohort are shown in Table 2.

**Table 2.**
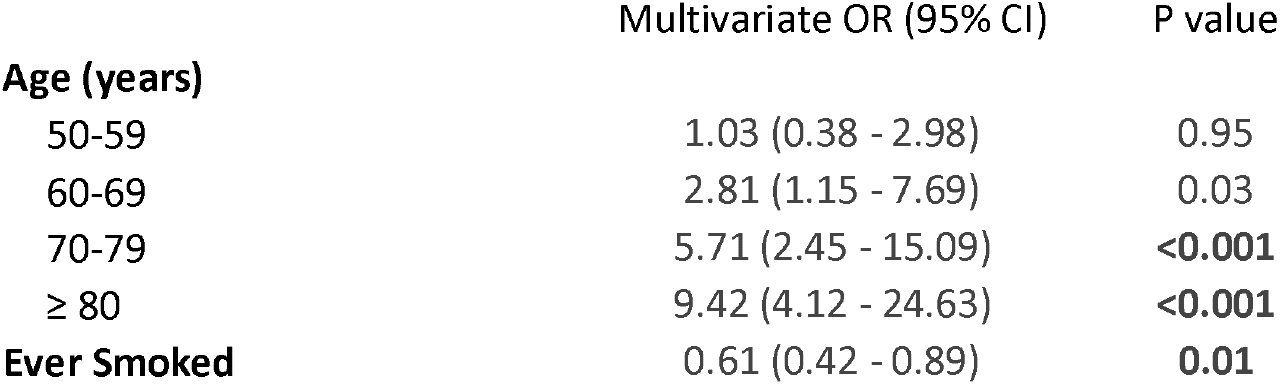
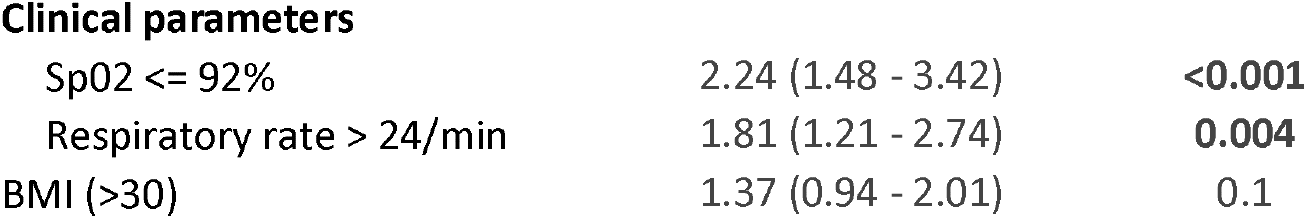
Clinical characteristics predicting mortality in second wave (PREDICT second wave cohort).

|  | Multivariate OR (95% CI) | P value |
| --- | --- | --- |
| **Age (years)** |  |  |
| 50-59 | 1.03 (0.38 - 2.98) | 0.95 |
| 60-69 | 2.81 (1.15 - 7.69) | 0.03 |
| 70-79 | 5.71 (2.45 - 15.09) | **<0.001** |
| ≥ 80 | 9.42 (4.12 - 24.63) | **<0.001** |
| **Ever Smoked** | 0.61 (0.42 - 0.89) | **0.01** |
| **Clinical parameters** |  |  |
| SpO2 ≤ 92% | 2.24 (1.48 - 3.42) | <0.001 |
| Respiratory rate > 24/min | 1.81 (1.21 - 2.74) | 0.004 |
| BMI (>30) | 1.37 (0.94 - 2.01) | 0.1 |

### Prospective validation of SOARS and 4C Mortality Score in PREDICT second wave cohort

In multivariate logistic regression, SOARS achieved an AUC for hospital mortality of 0.80 (Table 3) in the PREDICT second wave cohort (95% CI 0.77 –0.83, P < 0.0001) and demonstrated negative predictive power of 83%, with positive predictive power of 60.4%. Odds ratios of predicting mortality using SOARS were the greatest for age (between 1.98 and 23.19), followed by low oxygen saturations (2.48, 95% CI 1.77 – 3.49), BMI (1.70, 95% CI 1.23 – 2.36) and respiratory rate (1.72, 95% CI 1.23 – 2.40). Presence of cerebrovascular disease was not significant (OR 0.78, 95% CI 0.55 – 1.36). Figure 1 shows SOARS score odds ratio predicting mortality in PREDICT second wave cohort.

**Table 3.**
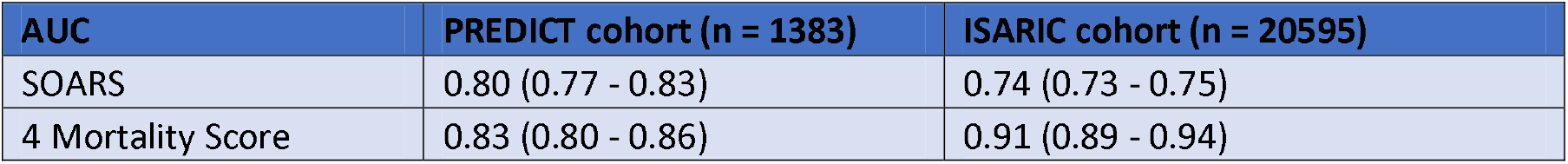
AUC for hospital mortality of scores in PREDICT and ISARIC second wave cohorts.

**Table 4.** SOARS and outcomes of patients in various treatment pathways utilised in wave 1 and 2: VH, ward, ward CPAP and ICU.

| Care Setting | Wave 1 | % | Wave 2 | % |
| --- | --- | --- | --- | --- |
| **VH and Discharged (n)** | 230/983 | 23.4% | 229/1383 | 16.6% |
| SOARS (Median (IQR)) | 3 (1 - 5) |  | 2 (0 - 2) |  |
| Re-admission | 25 | 10.9% | 27 | 11.8% |
| Mortality | 2 | 0.8% | 2 | 0.9% |
| **Ward (n)** | 632/983 | 64.3% | 958/1383 | 69.3% |
| SOARS (Median (IQR)) | 4 (2 - 5) |  | 4 (2 - 5) |  |
| 0-1 | 116 |  | 113 |  |
| 2 | 75 |  | 99 |  |
| 3 | 86 |  | 115 |  |
| Mortality | 212/632 | 33.5% | 223/958 | 23.3% |
| **Ward with CPAP (n)** | 41/983 | 4.2% | 17/1383 | 1.2% |
| SOARS (Median (IQR)) | 2 (1 - 4) |  | 3 (3 - 6) |  |
| Mortality | 20 |  | 2 |  |
| **ICU (n)** | 87/983 | 8.8% | 179/1383 | 12.9% |
| SOARS (Median (IQR)) | 3 (2 - 5) |  | 4 (3-5) |  |
| Mortality | 54/87 | 62.1% | 77/179 | 43% |

**Figure 1.**
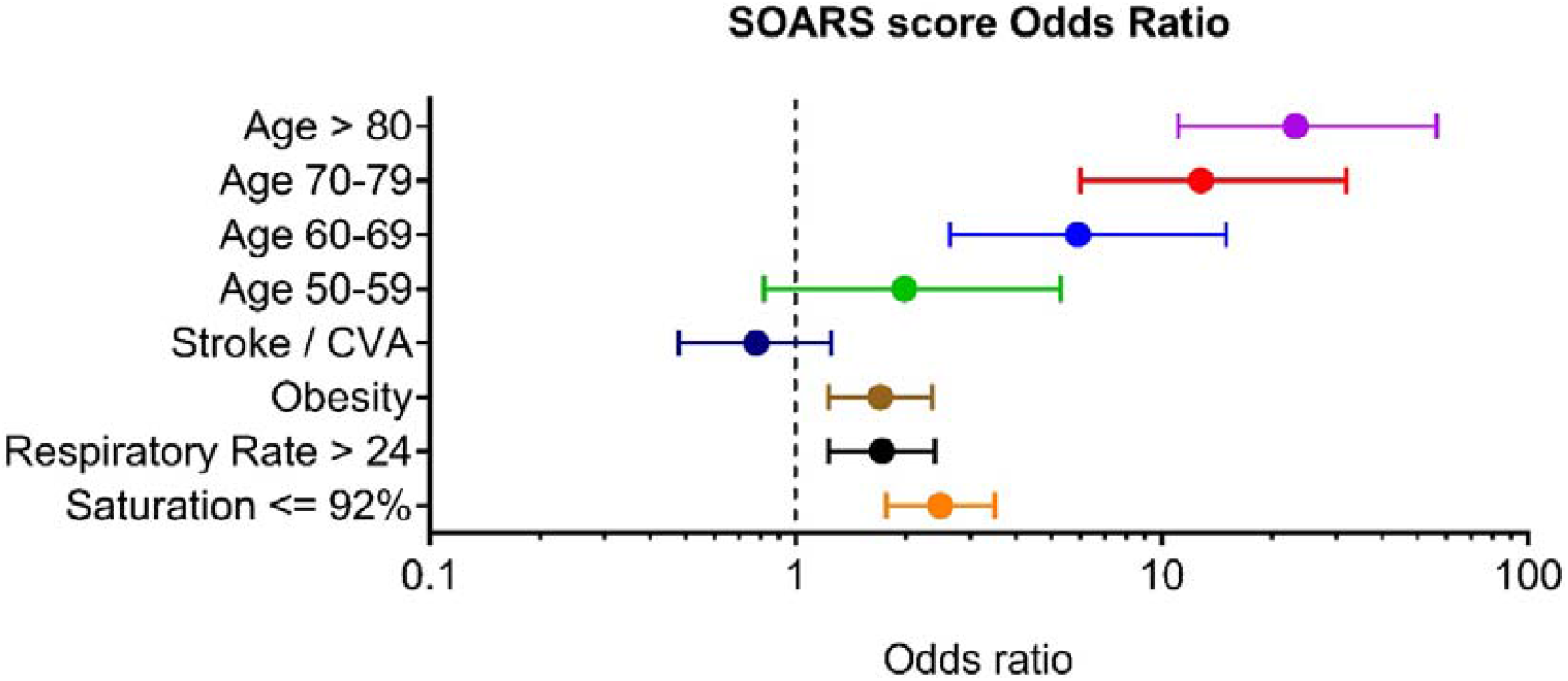
SOARS odds ratio predicting mortality in PREDICT second wave cohort

SOARS applied to the PREDICT second wave cohort had a linear relationship between score severity and in-hospital mortality (Figure 2). Of note, no deaths were noted in the subgroup who scored 0 (n = 101). Compared to the first wave, mortality significantly reduced in the second wave especially in the more severe disease, scores between 3 and 8, with a mortality reduction between 10% and 21%. Performance of the 4C Mortality Score in the PREDICT second wave cohort also showed increasing mortality with score severity, but less linearly as seen in SOARS (Figure 2).

**Figure 2.**
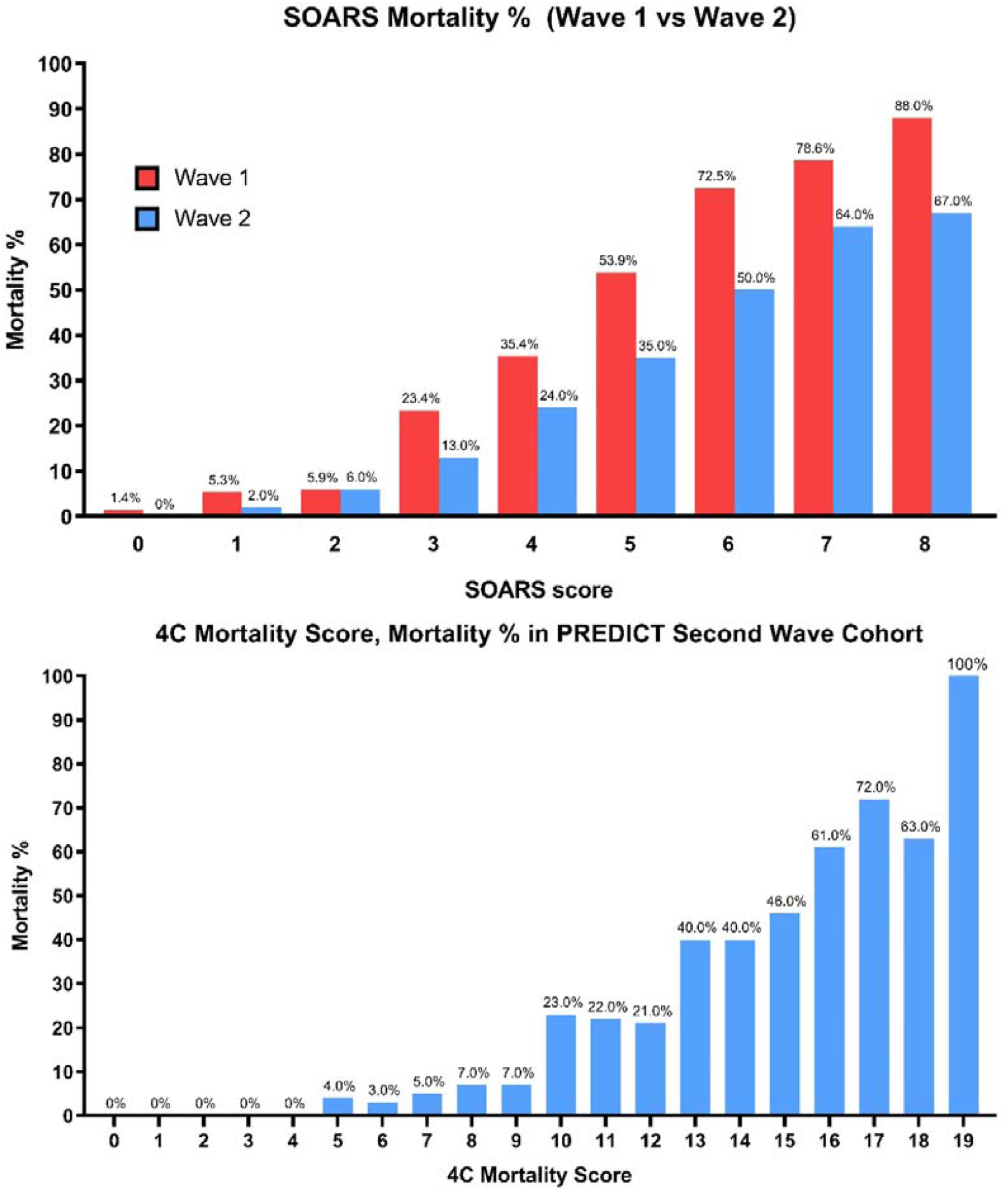
SOARS and 4C Mortality Score mortality in first and second wave PREDICT cohort

### External validation of SOARS and 4C Mortality Score on ISARIC second wave data

We looked at performance of scores derived from the first wave in an additional 20,595 UK second wave data provided by ISARIC (Figure 3). Again, the SOARS score showed a linear relationship with mortality until a score of 6 which then tapers in the higher scores. The 4C Mortality Score showed score severity correlating with mortality in the ISARIC second wave cohort, with similar safety to SOARS for the low risk (less than a score of 7). AUC of SOARS and 4C Mortality Score looking at sensitivity and specificity of scores in both the PREDICT and ISARIC second wave cohorts predicting mortality is shown in Table 3.

**Figure 3.**
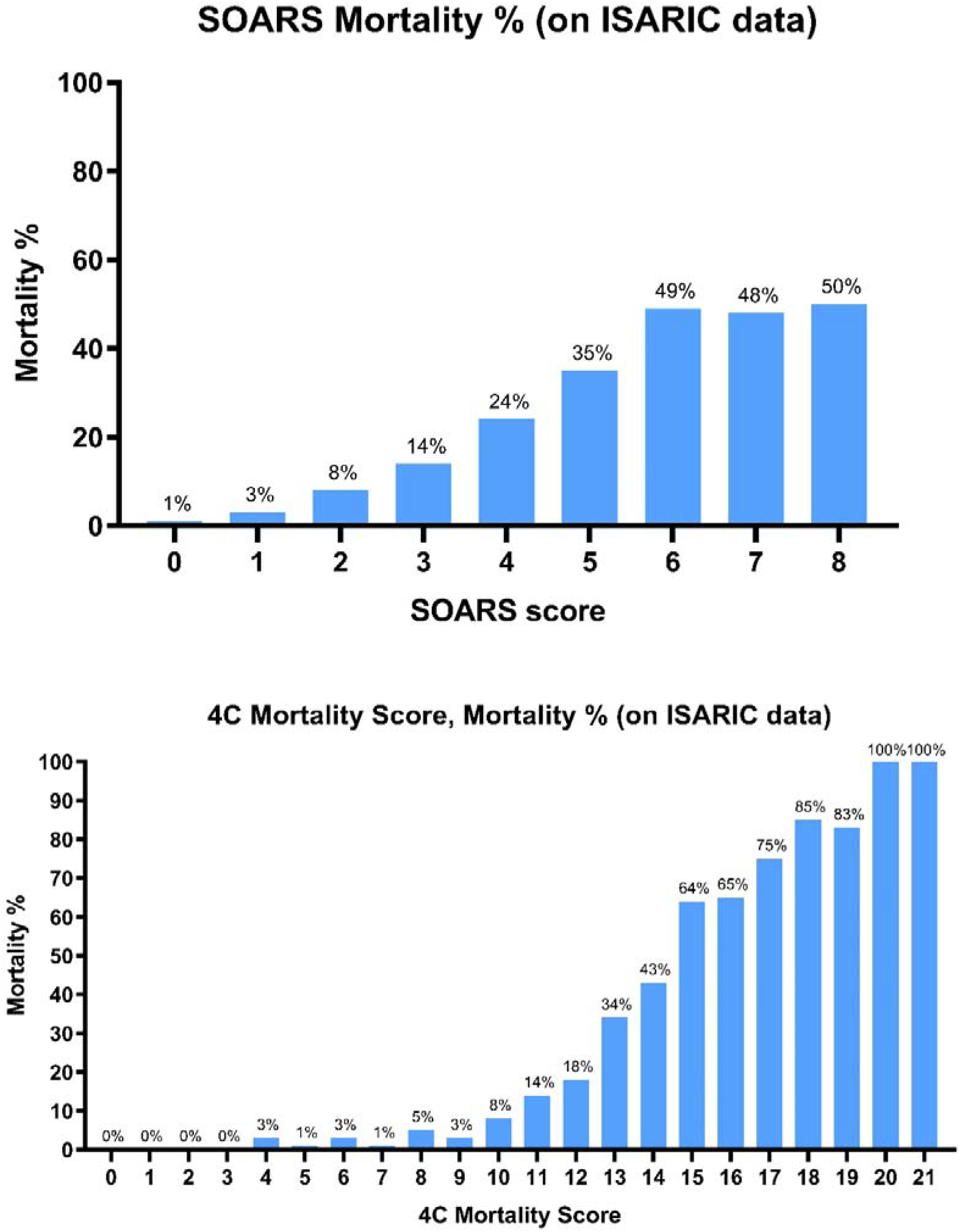
SOARS and 4C Mortality Score in the ISARIC second wave cohort

### Virtual Hospital

16.6% (n = 229/1383) of patients in the PREDICT second wave cohort with SOARS scores of 0 or 1 on presentation were discharged to the VH, where only 11.8% (27/229) of these patients required subsequently hospital admission and 0.9% (2/229) mortality. 8.1% (113/1383) of the PREDICT second wave cohort were admitted despite scoring SOARS 0 or 1, with mortality of 2.6%. 9.5% (309/3257) of patients in the ISARIC second wave cohort were admitted with SOARS of 0 or 1, with a mortality of 2%.

## DISCUSSION

The COVID-19 pandemic continues to escalate and evolve globally with lethal consequences, especially in South Asia and Latin America, despite remarkable advances in treatment, vaccination, and public health measures^22,28-30^. This may reflect increased infectivity and virulence of SARS-CoV-2 variants and inadequate healthcare response^4,6,7,15,20,21^. Infection may in time be adequately managed by vaccination and prudent public health measures, but this needs to be considered in tandem with the more urgent priority of ensuring appropriate hospital resource allocation for the acutely unwell while providing normal healthcare to the rest of the population^1,2,31-33^. We report prospective validation of two published scores derived from the COVID-19 first wave, in the evolving pandemic: SOARS, a clinical assessment-based triage score, and 4C Mortality Score, a hospital-based mortality score which is dependent on both clinical and investigation results^24,25^. We confirm the ongoing utility of SOARS for discharge in a multi-site UK study for score 0 and 1. The 4C Mortality Score retains utility for hospitalized patients for safe discharge below a score of 7.

As lockdown is lifted and routine life resumes in the United Kingdom, with high vaccination rates in the over 50s, preparation for a new rise of SARS-CoV-2 cases in a less comorbid population is paramount^26^. Learning from the pandemic, taught us that preserving limited healthcare resources is crucial to enable continued normal services in tandem with the needs of the pandemic^1^. The older and comorbid population have accrued morbidity, both from the pandemic itself and the absence of regular complex care needs, therefore stressing the importance of resuming regular healthcare services^1,31,32^. The prospective validation of the SOARS score, based only on clinical assessment as a mortality and discharge score, permits risk-assessed discharges to both the ‘virtual hospital’ (VH) and community care, reducing burden on hospitals. More importantly, the SOARS score without diagnostic investigations, is as valid prospectively in the second wave as resource intensive scores, making it a valuable tool for study and use in pandemic peaks in low resource systems.

The accelerated use of telemedicine in many countries for the pandemic enabled low-risk patients to be followed up in a VH in the community. VH has successfully been used by Atrium Health, a US integrated healthcare organization where low-risk patients were defined by the modified DSCRB-65 of less than 2^34^. Of the 1293 patients they followed-up in VH, only 40 (3%) required hospitalization, where in the re-admitted cohort 18% required ventilator support and 5% mortality during their hospital admission^34^. Our experience using a VH model with integrated distant monitoring during the first wave, where 900 low-risk patients (defined by a NEWS score < 2 and CRP< 50) were followed-up, resulted in 8.1% re-admission and 2% mortality^27^. The initial success of the VH model stimulated the need for a safe discharge scoring system, where the SOARS score was developed. SOARS scores of 0 and 1 were used to discharge and follow-up patients in the VH in a reliable, safe, and easy to use pathway, with integrated mortality at the various scores enabling educated decision making between clinician and patient (Supplement 2). Using the SOARS in triaging discharge during the second wave, 16.6% (229/1383) of patients were discharged home or to the virtual hospital, where 11.8% (27/229) of those discharged were re-admitted and 0.9% mortality. However, the use of SOARS score is still suboptimal as 8.1% (113/1383) were still admitted despite having scores of 0 or 1. Applying the SOARS score nationally in the ISARIC second wave cohort could have potentially avoided admission in an additional 9.5% (309/3257) of patients. We therefore recommend using the SOARS score as an integral part of managing SARS-CoV-2 cases as shown in Figure 4.

**Figure 4.**
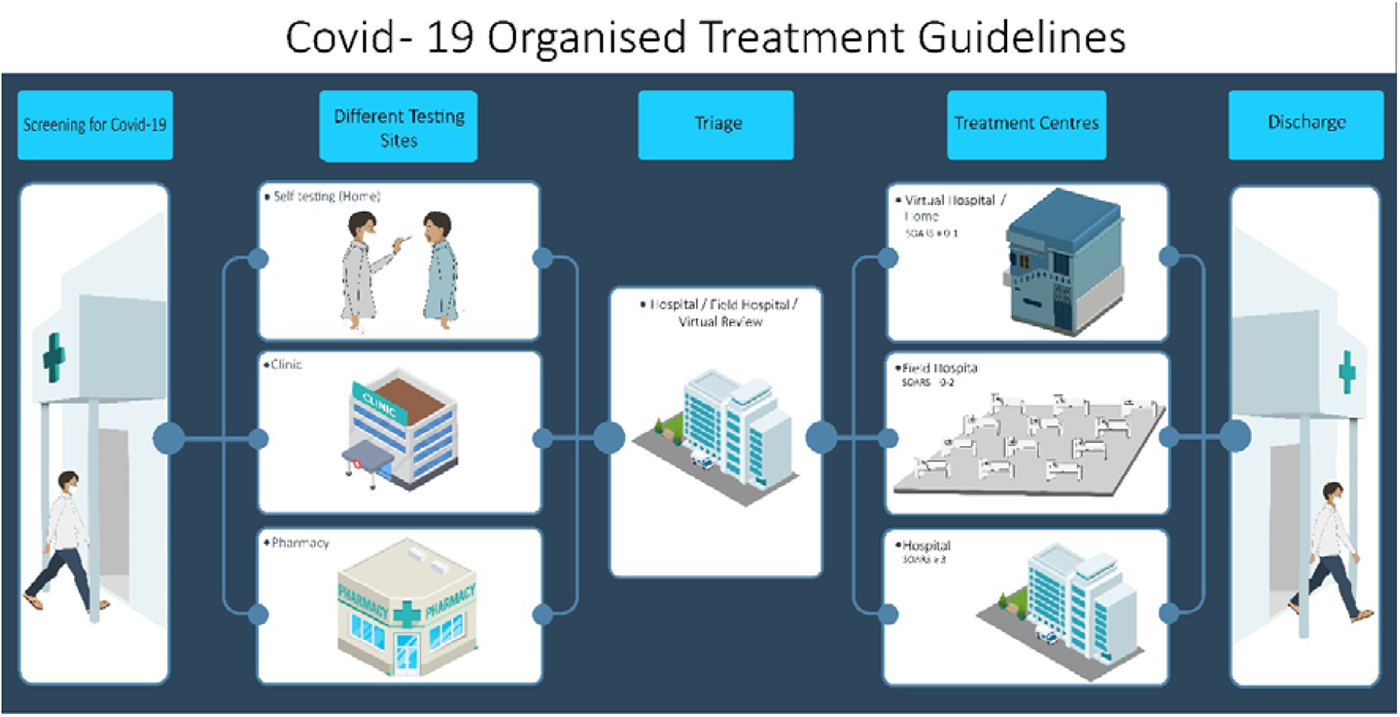
Using SOARS in COVID-19 management

Improved public health measures and lower virulence of SARS-CoV-2 may account for the reduced admissions from care homes with frailty, dementia, and stroke (CVA) in the PREDICT second wave cohort. It is unlikely to reflect treatment advances, as these patients were less seen or admitted and mortality reducing treatments were only provided to those hospitalized. Vaccinations were unlikely to play a role as were only started in the older population in Hertfordshire at the end of December 2020 and beginning of January 2021. The improvement in mortality in the VH population who did not receive Dexamethasone suggests some reduction in SARS-CoV-2 virulence in the second wave. It is of interest to note the loss in significance of some variables that accounted for mortality in the first wave, including smoking, obesity, CVA, and lymphopenia. A number of these comorbidities are associated with inflammation, particularly smoking, which may have been attenuated with the early use of Dexamethasone.

One of the main limitations of the SOARS score is its dependance on age. Older patients automatically trigger a higher mortality score regardless of their comorbidities and clinical presentation, a limitation in all COVID-19 severity scores due to high weighting for age. This may change with vaccination and will require review in post-vaccination waves. If viral subtypes evade vaccination or boosters are delayed, age may still be relevant to scoring^7^. Additionally, multivariate analysis from the second wave revealed that pre-morbid CVA (OR 0.78), despite affecting a similar proportion of SARS-CoV-2 cases in both waves (Table 1), was no longer associated with mortality, an observation difficult to explain. Another limitation of this study is the absence of SARS-CoV-2 serotyping of individual patients which would help assess the contribution of B.1.1.7 variant in presentations and outcomes. This may have provided some clarity to the differential contribution of the variant to treatment, public health measures and host characteristics resulting in the reduced mortality in the second wave. The contribution of Remdesivir and Tocilizumab were also not discussed in view of the small number of patients treated with these agents and as it was not the primary objective of this study. Effects of vaccination did not contribute significantly to the second wave, a limitation that requires further review in future.

SOARS, a clinical score developed to enable safe discharge to the community and VH, now prospectively validated in both the derivation and multi-site UK cohorts, remains relevant to its purpose despite the evolution of the virus and the pandemic. This score developed in the parent variant remains relevant despite significant displacement by the B.1.1.7 variant, a prominent subtype in the second wave^16,17^. The relevance of this score in overwhelmed healthcare systems allows expedient, safe and reliable decision making. Utility of the SOARS score in the South African, Brazilian and the Indian SARS-CoV-2 variants warrants further study. Further studies in future waves should continue to be performed to ensure the tool’s validity in purpose and safety in SARS-CoV-2 infection.

## SUMMARY BOX

What is already known on this topic:

- COVID-19 prognostication scores are all based on first wave of the pandemic; however, the pandemic is evolving due to SARS-CoV-2 subtypes, socio-economic healthcare responses and/or different host-viral interactions raising the need for prospective validation of existing scores.
- The SOARS score, a clinical assessment-based triage score, and 4C Mortality Score, a hospital-based mortality score which is dependent on both clinical and investigation results are both validated prognostication scores derived in the first wave.
- Given the uncertainty of SOARS and 4C Mortality Score performance in the evolving pandemic, considerable interest exists in showing these tools’ validity in purpose and safety in SARS-CoV-2 infection.

What this study adds

- Prospective validation in a large single-site (PREDICT) and multi-site (ISARIC) UK cohorts determined relevance of the SOARS and 4C Mortality Score despite the evolving COVID-19 pandemic with treatment advances and SARS-CoV-2 variants.
- The SOARS score is a safe, easy to use and reliable triage score enabling rapid discharge, providing appropriate allocation of hospital resources to the need of the pandemic whilst enabling resumption of normal delivery of healthcare.

## Data Availability

Data are available upon reasonable request. Deidentified participant data may be requested from the corresponding author following publication of the study.

## Contributors

HG, AN and RV contributed equally. HG and AN are joint first authors. RV conceived the study and designed with input from HG, AN, NN, AB. HG, AN and RV supervised various aspects during conduct of the study. HG, AN, HM, WE, BC, TV, MSc and RV contributed to data acquisition. AN, BC and MSh analysed data which was interpreted by all authors. All authors approved the final version to be published and agree to be accountable for all aspects of the work. HG and AN are the guarantors: accept full responsibility for the work and/or the conduct of the study, had access to the data, controlled the decision to publish and affirm that the manuscript is an honest, accurate, and transparent account of the study being reported and that no important aspects of the study have been omitted. The corresponding author attests that all listed authors meet authorship criteria and that no others meeting the criteria have been omitted.

## Aknowledgement

We thank Abdul-Muiz Azri Yahya, Damilola Longe, Eleanor Croft, Hee La Lee, Iman Darwish, Horiyo Nur, Jishanthan Ragunathan, Kholiwe Kutshwa, Meghna Prabhakar, Mihira Patel, Nabiah Malik, Nafissa Hussain, Sceyon Mohan, Shamira Ghouse and Simon Saldanha, doctors at West Hertfordshire Hospitals NHS Trust for their assistance in this study. We also thank the International Severe Acute Respiratory and Emerging Infections Consortium (ISARIC) for providing us with the multi-site data.

## Patient consent for publication

Not required.

## Ethics approval

Ethical approval was provided by Stanmore Research Ethics Committee, London, England (IRAS ID: 283888). The study is registered by the National Health Service Health Research Authority under the reference 20/HRA/2344.

