## Supplement 1 for "Relevance of prediction scores derived from the SARS-CoV-2 first wave, in the UK COVID-19 second wave, for early discharge, severity and mortality: a PREDICT COVID UK prospective observational cohort study"

### Appendix. Pre-specified study protocol for clinical data collection

**PREDICT COVID Clinical Data Collection protocol**
**Date: 1st March 2020**

---

Aims
1. To prospectively collect data on all adults (> 18 years) with laboratory-confirmed SARS-CoV-2 infection (COVID-19) presenting to Watford Hospital, West Hertfordshire NHS Trust, during the first wave of the SARS-CoV-2 pandemic, including an outcome of in-hospital death or hospital discharge,
2. To develop a prognostic (risk prediction) score using the above derivation data,
3. To construct a practical clinical scoring system for predicting mortality (and risk of morbidity) after validation of score against external, i.e. independent cohorts of COVID-19 patients from other UK sites,
4. To include and assess outcomes of patients referred to the COVID-19 Virtual Hospital (out-of-hospital monitoring) in the same NHS Trust,
5. To monitor and characterise surviving patients for up to 12 months from the time of confirmation of SARS-CoV-2 infection, including the domains of psychological, physiological and radiological impairment and recovery.

Primary outcome
In-hospital mortality with minimum 30-day follow-up data.

Secondary outcomes
Longer term mortality and morbidity: radiological, psychological and cardiorespiratory. This will also be assessed against ongoing health care needs.

Patient inclusion
All adult patients (aged >18 years) with SARS-CoV-2 real-time reverse transcriptase polymerase chain reaction (rRT-PCR) confirmation.
Completed admission and outcomes at 3 months and 12 months.

Inclusion criteria:
- Readily available patient or clinical characteristic to attending clinicians upon presentation to hospital (Accident & Emergency department, Acute Medical Receiving Unit)
- Blood markers should be commonly measured and results available for review within the first 24 hours of admission
- All parameters relating to oxygen supplementation and advanced respiratory support including one or more of: continuous positive airway pressure (CPAP), bilevel non-invasive ventilation (NIV), high-flow nasal cannula (HFNC) oxygenation and invasive mechanical ventilation (IMV) needs

Exclusion criteria
All SARS-CoV-2 rRT-PCR negative patients irrespective of clinical suspicion of COVID-19.
All individuals aged <18y.

Selection of candidate variables for initial data collection
Candidate variables were chosen based on knowledge of their potential association/s with SARS-CoV-2 infection and clinical disease (COVID-19). A systematic literature search was undertaken to identify these variables with respect to their predictive association for mortality and other adverse outcomes including COVID-19 severity and disease-related complications such as requirement for critical care and the development of COVID-19-associated acute respiratory distress syndrome (ARDS).

Systematic literature for English language articles in the following search databases: PubMed, EMBASE, WHO Medicus, Web of Science and Google Scholar (particularly for pre-print publications on medRxiv). Search terms included SARS-CoV-2; COVID-19; coronavirus; ARDS; pneumonia; sepsis; influenza; risk prediction; risk score; prognosis; validation. No date restrictions were imposed.

Statistical analysis for derivation and validation modelling

In analysing the data collected from the derivation (West Herts) cohort, categorical variables will expressed as frequency (%), with significance determined by the Chi-squared test. Continuous variables will be analysed for median (interquartile range) or mean (standard deviation) outcomes and analysed by the t-test, Kruskal-Wallis or Mann-Whitney U test, as appropriate. Missing data in the derivation cohort will be expected even with prospective data collection; missingness of data will be assumed to be at random and handled by multiple imputation by chained equations (MICE) with at least ten imputations, provided the proportion of missingness for the defined parameter constitutes no more than 20% of the cohort. Collated data will be subjected to univariate and multivariate logistic regression in order to determine odds ratios (OR) for in-hospital mortality. The latter will also be internally validated by bootstrapping using a minimum of 1000 re-samples. Predictor interactions will be analysed by appropriate methodology such as the likelihood ratio (LR) test comparing broad and narrow (constrained) models.

All performance metrics against external validation cohorts will be analysed using the prediction score and not with the multivariate regression model. The performance of the derivation model will be assessed for discriminatory ability (area under the receiver operating characteristic, AUROC) and calibration (graphical representation of Hosmer-Lemeshow analysis).

All statistical analyses including risk modelling calculations will be performed using STATA, version 16 (Stata Corp., Texas, USA).

*R Vancheeswaran, F Chua, A Draper, T Vaghela and A Barlow (study design and responsible persons for data analysis and interpretation)*

*MARCH 2020*
