## Supplement 2 for "Relevance of prediction scores derived from the SARS-CoV-2 first wave, in the UK COVID-19 second wave, for early discharge, severity and mortality: a PREDICT COVID UK prospective observational cohort study"

Patient sticker

### COVID-19 PATHWAY
**For adult patients ≥16 years over (Suspected or Confirmed)**

West Hertfordshire Hospitals NHS Trust

#### INITIAL ASSESSMENT

**Date/Time of assessment:** _____/_____/_____ ____:____

**Name of clinician:** ______________________________

Covid-like symptoms (Fever, myalgia, persistent cough, dyspnea, change/loss of smell or taste)

**NO** → Step down from Covid zone after discussion with senior

**YES** ↓

1. Send Covid-19 PCR Swab ☐
2. Send Flu swab ☐
3. **Calculate SOARS score** →

**SOARS Score (Please circle)**

| | |
|---|---|
| **$SpO_2$:** | **<92% (1 point)** |
| **Obesity:** | **Yes (1 point)** |
| **Age:** | **50-59 (1 point)** |
| | **60-69 (2 points)** |
| | **70-79 (3 points)** |
| | **>80 (4 points)** |
| **RR:** | **>24 (1 point)** |
| **Stroke:** | **History (1 point)** |

**Total: SOARS = ____________**

**0-1** | **2** | **3 OR ABOVE**

##### GREEN — SOARS SCORE 0-1

1) **Discharge to Virtual Hospital****
2) **Give patient Advice Pack (with oximeter)**

****Make referral via Infloflex Web COVID 19 or if Infoflex unavailable**

##### HIGH RISK CRITERIA

| | |
|---|---|
| Age: | <50 years |
| | AND |
| R/R: | >24 |
| SPO2: | <92% RA |
| NEWS: | >2 |

##### ORANGE — SOARS SCORE 2

**Is the patient High Risk? (Check high risk criteria)**

**NO** → **Complete treatment in GREEN BOX**

**YES** ↓

1) **ADMIT UNDER MEDICS**
2) **Send bloods (Set 1)**
3) **Book Chest X-Ray**
4) **Calculate COVID LONG SCORE**

**If CLS <7**

5) **Assess oxygen requirements**
6) **Prone if <94% Room Air**
7) **Aim for ≥94% (unless COPD or known Type II RF)**

**If CLS ≥7**

5) **Complete treatment in RED BOX**

##### RED — SOARS SCORE ≥ 3 or COVID LONG SCORE ≥ 7

1) **ADMIT UNDER MEDICS**
2) **Send bloods (Set 1 and 2)**
3) **Book Chest X-Ray**
4) **IV fluids if indicated**
5) **Antibiotics if PCT >0.5 (refer to Microguide)**
6) **Aim for ≥94% (unless COPD or known Type II RF)**
7) **Calculate Rockwood Score (CFS)**
8) **Access DNAPCR and TEP**

**COVID LONG SCORE (Please circle)**

| | |
|---|---|
| **Ever smoker:** | **Yes (1 point)** |
| **Dementia:** | **Yes (1 point)** |
| **CKD:** | **Stage 1 (1 point)** |
| | **Stage 2 (2 points)** |
| | **Stage 3 (3 points)** |
| | **Stage 4 (4 points)** |
| | **Stage 5 (5 points)** |
| **WCC:** | **>11 (1 point)** |
| **Lymph:** | **<0.7 (1 point)** |
| **CXR:** | **>4 zones (1 point)** |

**Total: SOARS + CLS = ____________**

**Bloods:**

**(Set 1) FBC; U/Es; CRP; LFT**

**(Set 2) Ferritin; D-Dimer; Procalcitonin; BNP; LDH; Troponin**

| SOARS score mortality | | | | | | | | |
|---|---|---|---|---|---|---|---|---|
| 0 | 1 | 2 | 3 | 4 | 5 | 6 | 7 | 8 |
| 1.4% | 5.3% | 5.9% | 23.4% | 35.4% | 53.9% | 72.5% | 78.6% | >78.6% |

### INPATIENT MANAGEMENT

**Time/Date of initial treatment: _____/_____/____ ____:____ Name of clinician: ____________________**

#### 1. INITIAL TREATMENT

*If RA SpO2<92% (unless COPD/OHS/CCF)*

- ➔ Give supplemental oxygen – target Sats up to 94%
- ➔ Prone if needed
- ➔ Anticoagulation as per risk of VTE: Therapeutic/Prophylactic (see Anticoagulation and Thromboprophylaxis policy). CTPA to be considered after consultant/respiratory review
- ➔ Perform PCT and **STOP** antibiotics if below 0.25 mcg/L (Refer to MicroGuide)
    1. Send sputum sample and blood cultures
    2. Choice of antibiotics based on hospital guidelines (community acquired pneumonia; hospital acquired pneumonia; sepsis)
    3. Total duration of antibiotics - 5 days
- ➔ Treat other comorbidities
- ➔ Prescribe Covid medications if indicated (see medications on page 4)

#### 2. CALCULATE CLINICAL FRAILTY SCORE (CFS) *(PLEASE CIRCLE)*

*Your assessment should be based on the patient's functional status TWO WEEKS prior to hospitilisation*

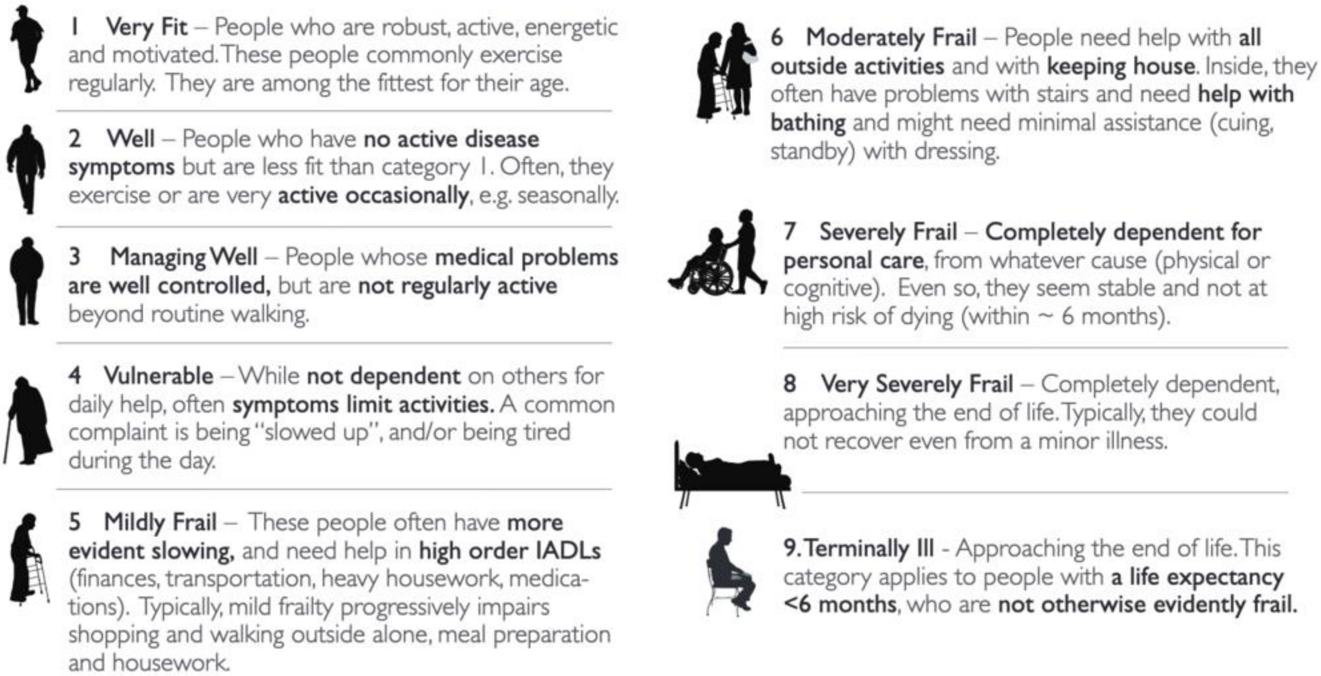


#### 3. MAKE ESCALATION DECISION AND DNACPR IF APPROPRIATE

| Please tick Escalation Decision chosen ☐ | Please tick Escalation Decision chosen ☐ |
|---|---|
| ***If CFS <5*** <br> → **FULL ESCALATION** | ***If CFS >=5*** <br> → **WARD BASED CEILING OF CARE** <br> ○ Complete TEP with senior clinician <br> ○ **Patients are not for ICU or NIV** |

### FULL ESCALATION

- ***If $SpO_2$ < 94% on $FiO_2$ of 0.6***
  *or CPAP > 10cm$H_2O$*
  *or PF ratio < 100 (PF ratio = arterial $PaO_2$ / $FiO_2$)*
  *or worsening at 24h/daily review*
  ➔ **Rapid assessment for intubation & transfer to ITU if appropriate**

- ***If $SpO_2$ < 94% on $FiO_2$ of 0.4***
  ➔ **Discuss with Respiratory Consultant (available 24hours**) if candidate for NIV and recruitment into Recovery Respiratory Support as shown here:

| ***Recovery Respiratory Support*** | | |
|---|---|---|
| **HIGH FLOW NASAL OXYGEN (HFNO)** | **CONTINUOUS POSITIVE AIRWAY PRESSURE (CPAP)** | **STANDARD CARE** |
| Start at 60L/min flow @ $FiO_2$ 0.6 | Start at 8 cm$H_2O$ @ FiO2 0.5 | Based on clinical assessment |
| Up titrate by increasing $FiO_2$ | Up titrate by increasing pressure by 2 or increasing FiO2 | |
| *Weaning*<br>Trial on Venturi face mask, if fails, back to HFNC | *Weaning*<br>If $FiO_2$<0.5 swap to A40 in CPAP mode with entrained $O_2$ to wean<br>Match settings when swapped over<br>Respiratory team review in one hour | |
| Assess daily clinical review; PF ratio; ABG; if deteriorating, discuss with Respiratory Consultant or ITU | | |

- ***If $SpO_2$ > 94% on $FiO_2$ of 0.4***
  ➔ **Standard care in the ward**

### WARD BASED CEILING OF CARE

- Always titrate treatment to maintain $SpO_2$ up to 94% (unless COPD/OHS/CCF where target is 88-92%)
- Therapeutic anticoagulation in ill patients with high suspicion for PE/DVT

***Not reaching $SpO_2$ up to 94% on $FiO_2$ of 0.4?***

➔ Discuss with Respiratory Consultant in consideration of HFNO: Start at 60L/min flow @ $FiO_2$ 0.6 or CPAP (8 cm$H_2O$ @ FiO2 0.5)

***If PF ratio <100 or declining oxygenation or high risk score***

➔ Timely discussion with family to enable palliative support

### MEDICATIONS – ACTIVE TREATMENT

*For more information about Covid-19 medications, contact Medicines Information.*

| **DEXAMETHASONE** | |
|---|---|
| **Indication** | **Treatment of Covid-19 on supplemental oxygen** |
| **Dose** | • 6mg dexamethasone orally or 6.6mg dexamethasone IV, once a day, 7-10 days<br>• If on IV, switch to oral dexamethasone as soon as possible<br>• Dexamethasone should be stopped on discharge unless patient is hypoxic or still symptomatic and not at baseline, in which case the treatment course should be completed post discharge<br>• When prescribing dexamethasone consideration needs to be given to the gastric ulcer protection effect of proton pump inhibitors according to local hospital policy |

**The following medications have inconclusive evidence – and should only be used with Resp/ITU/Micro Consultant recommendation**

| | **REMDESIVIR** | **TOCILIZUMAB** (discuss with Resp./ITU consultant) |
|---|---|---|
| **Indication** | • **Covid-19 confirmed, within acute viremic phase** (after 7 days of symptom start)<br>• **Lymphopenic**<br>• **Need for oxygenation but not requiring mechanical ventilation**<br>• Ensure eGFR >30; body weight >40kg, and ALT <200 | • **High dose methylprednisolone sparing agent targeting cytokine storm in <u>critically ill</u> <u>confirmed COVID-19</u> patients at presentation with fever >38 and Type 1 Respiratory failure due to pneumonia.**<br>• **And ONE or more of the following high-severity markers:**<br>○ CRP >50/Ferritin >500μg/L / D-dimer >1000mg/ml / LDH >250U/L<br>**Exclusions:**<br>○ Known hypersensitivity to Tocilizumab<br>○ Co-existing infection which may be worsened by Tocilizumab<br>○ More than 24hrs on respiratory support<br>○ Evidence of immunosuppression<br>○ Platelets <50$\times10^9$/L, ALT >200, Neut. <2 $\times10^9$/L |
| **Dose** | Day 1 – single loading dose of Remdesivir 200 mg given by intravenous infusion<br>Day 2 onwards – 100 mg given once daily by IV infusion.<br>**<u>The total duration of treatment should be 5 days</u>**<br>Stop treatment if patient not on oxygen or discharged | • 8mg/kg (up to a max. of 100kg) by IV<br>• Should be diluted in 100mL bag of 0.9% Sodium Chloride after removing equivalent volume of 0.9% Sodium Chloride and given over 1 hour<br>• A single dose to be administered with option to repeat dose in 12-24 hours if initial dose has not caused sufficient improvement |

### MEDICATIONS – PALLIATIVE CARE

| **Indication** | **Drug** | **PRN S/C Dose** | **Syringe Driver (CSCI) over 24 hours** |
|---|---|---|---|
| Pain/Cough | Morphine Sulphate (half dose in elderly patients. If eGFR <30 use oxycodone) | 2.5-5mg 2-4 hourly | 10-20mg |
| Breathlessness | Midazolam + Morphine sulphate (as above) | 2.5-5mg 2-4 hourly | 10mg |
| Delirium | Haloperidol (half dose in elderly) | 1-5mg in 1-3 divided doses over 24h, max 5mg/day | N/A |
| Delirium *if end of life* | Levomepromazine | 25mg | 50mg |
| | Midazolam | 5mg 2-4 hourly | 15mg |
| Nausea and Vomiting | Cyclizine | 25mg 8 hourly | 100-150mg |
| Seizures | Midazolam | 5-10mg 2-4 hourly | 30-60mg |
| Respiratory secretions | Glycopyrronium | 0.2-0.4mg 4 hourly | 1.2-2.4mg |

**If symptoms are not adequately controlled, please contact the palliative care team on 01923 217930 or bleep 1006**

Version date: 29/01/2021 // Review date: 18/12/2021 // Version no: 5.1 // **Print Code: CovPath5.1**
Authors: R. Vancheeswaran, A. Navarra, S. Khan, N. Nordin, R. Mogal, A, Jayarathnam, A. Barlow., CPG Prog.
